# Ensemble model estimates of the global burden of measles morbidity and mortality from 2000 to 2019: a modeling study

**DOI:** 10.1101/2021.08.31.21262916

**Authors:** Heather Santos, Kirsten Eilertson, Brian Lambert, Sarah Hauryski, Minal Patel, Matthew Ferrari

## Abstract

**Background:** Measles remains a significant source of childhood morbidity and mortality worldwide. Two doses of measles containing vaccine are recommended for all children and delivered through a combination of routine and supplemental immunization activities. Uncertainty about the degree to which second dose opportunities reach previously unvaccinated children presents a challenge in the assessment of vaccination programs and the estimation of the global burden of measles disease and mortality.

**Methods:** We fit an ensemble of models that represent alternative assumptions about the degree to which second dose opportunities reach previously unvaccinated children to routine measles surveillance from 100 countries. Using maximum likelihood we selected the best fit model for each country. We compare the resulting estimates of the burden of measles disease and mortality to existing methods for estimating the burden of measles that assume that second dose opportunities are independent of receipt of the first dose.

**Findings:** We find that 78 of 100 countries are best-fit by a model that assumes that second doses that are delivered through supplemental campaigns are preferentially delivered to children who have received a first dose. Using a country-specific best-fit model we estimate that measles mortality has declined by 73% from 2000-2019 compared to an estimated decline of 83% using an assumption of independent doses in all countries.

**Interpretation:** Despite large decreases in measles cases over the last two decades, the observed trajectories in most countries suggest that supplemental immunization activities are disproportionately reaching previously vaccinated children. To accelerate measles reduction goals efforts to reach unvaccinated children through supplemental activities and second dose opportunities should be intensified.

**Funding:** Bill and Melinda Gates Foundation, World Health Organization

## Introduction

Despite large declines in measles incidence worldwide since 2000, measles cases have increased in all regions since 2017 (1). This increase has been in part due to stagnation of vaccination coverage (2) and the consequent resurgent outbreaks facilitated by the accumulation of the non-vaccinated population (3–5). As the majority of measles infections and deaths in the world go unrecorded, statistical estimates of these quantities are critical to evaluate vaccination program performance and progress towards control and elimination goals.

All methods to estimate the burden of unrecorded mortality come with potential biases and sources of uncertainty. Simons et al (6) proposed a method to estimate the burden of infection from country-level, annual reported measles cases. Though an annual model is coarse with respect to the fast dynamics of measles transmission and seasonality (7), this timescale allows a common model to be fit to all countries over a long time frame (1980 to present) based on reporting by member states to the World Health Organization (WHO) to permit consistent evaluation of changes in global measles burden and the impact of vaccination programs. Here we describe an update to the original Simons et al(6) methodology to address four simplifying assumptions of the original methodology.

First, the original Simons model assumed a phenomenological relationship between the measles attack rate and the proportion of the population that is susceptible that was linear when susceptibility was low. Thus, while the probability that a susceptible individual is infected is smallest when the proportion susceptible is small, it increases most quickly as the proportion susceptible increases at small values. This is inconsistent with the expected non-linear behavior of indirect protection below the herd immunity threshold (e.g. 3-5% of the population susceptible)(8) where the attack rate should increase slowly as the susceptible population increases below the threshold, and sharply thereafter. Here we include this non-linear effect.

Second, the original Simons model assumed that the error distribution around burden estimates was Gaussian with constant variance. This assumption allowed fast computation, but leads to counterintuitive phenomena; specifically, the width of confidence bounds do not scale with burden and negative lower bounds are permitted (though were excluded post hoc). Here we employ a binomial error distribution that scales with burden and is constrained to positive values.

Third, the original Simons model was not explicitly age-specific; the age distribution of the susceptible population, and thus impact of age-targeted supplemental immunization activities (SIAs) was approximated based on region and measles vaccination coverage (6). Here we extend the model to explicitly account for the age distribution of the susceptible population, which allows direct calculation of the impact of SIAs on individual targeted age groups.

Last, the original Simons model assumed that all measles containing vaccine doses were delivered independently of prior vaccination. Though the explicit correlation between vaccination doses is difficult to know for all countries, numerous studies have shown that second dose opportunities (through routine or SIA doses) tend to underperform nominal coverage values because of inequities in access to care (9–11); i.e. second opportunities are disproportionately delivered to those who received a first dose. To address this uncertainty in the interaction among vaccine doses, we fit an ensemble of models to each country that reflect alternative assumptions about the correlation between routine first dose, routine second dose, and SIA doses. Thus, rather than making a single assumption for all countries, we use maximum likelihood to select the country-specific evidence-based model for the correlation among vaccine doses.

Below, we expand the details of these model updates and compare the resulting estimates of the global burden of measles incidence and mortality from 2000 to 2019. We then report on the insights gained from fitting alternative measles dose correlation models and the implications for projecting the future impact of measles vaccination investments.

## Methods

### Data

Annual measles cases and vaccination coverage with the first and second dose of routine measles containing vaccine (MCV) from 1980 to present were taken from the WHO/UNICEF Joint Reporting Form (12). Reported measles cases and vaccination coverage are reported annually by member states to WHO and the United Nations Children’s Fund (UNICEF). Using coverage data as well as vaccination coverage survey data sourced from published and grey literature, WHO and UNICEF estimate the national immunization coverage for timely fist dose of MCV (MCV1) and second dose of MCV (MCV2) administered through routine immunization services (not mass campaigns), updating the entire time series from 1980-previous year (13–15). For MCV1, if MCV1 is given at age <1 year, the denominator is among children aged 1 year or, if MCV1 is given at age ≥1 year, among children aged 24 months. Children who receive late doses past these ages are not included in the calculation. For MCV2, it is calculated among one birth cohort of children at the recommended age for administration of MCV2, per the national immunization schedule. Again, vaccination given after the recommended age is not counted towards the estimated MCV2 coverage.

A list of SIAs conducted between 1980-2019 was provided by WHO based on reporting to WHO-UNICEF annually by member states (16). SIAs are frequently mass vaccination campaigns that attempt to vaccinate all persons within a target age group in a few days or weeks. The list included the upper and lower age target and coverage achieved for each campaign. For age limits that were not in integer years, we scaled the coverage relative to the portion of the age cohort covered: e.g. a lower age target of 9 months vaccinated is calculated as .25 x coverage in the 0-1 age cohort. The coverage of subnational SIAs was scaled to the proportion of the total population included in the subnational target. The date of SIAs was coded only as the year in which it was conducted. Uncertainty about whether the impact of SIAs is first observed in the year of the campaign (i.e. campaign was conducted early in the year) or in the year following the campaign (i.e. campaign was conducted late in the year) was addressed by fitting alternative models (see below).

Population data on live births and total population in each year came from the United Nations World Population Prospects 2019 (17).

Surveillance identifies a small proportion of cases that occur, even in countries with very good surveillance—for example some persons will never seek care and thus remain invisible to the surveillance system, while other times clinicians fail to report cases to public health. However, during an outbreak, surveillance is frequently intensified as recommended by WHO (18), with awareness raised among the public and clinicians. This can lead to improved reporting during this period. To account for these periods of increased reported, WHO experts review the surveillance data to determine if the country is having a high reporting rate in any given year. A list of years when reporting was enhanced in individual countries, e.g. due to outbreak response, was provided by WHO.

### Model description

Similar to the approach described in Simons et al (6), we present a 2-stage approach to estimating the burden of measles mortality. In the first stage we estimate the burden of measles infection using a state-space model. In the second stage, we estimate the burden of measles mortality by applying age-specific case fatality probabilities to the estimates from the first stage.

The model and fitting algorithm are described in Eilertson et al (19). Here we provide a brief review and description of changes. The populations of susceptible,, and infected, *I*_*t*_, individuals at year, *t*, are represented as vectors of length A = 100, where the elements 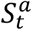 and 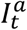 are the number susceptible and infected in year *t* in age one-year age cohort *a*. Individuals are born with maternal immunity, then after a period of 6 months become susceptible. At each year individuals either 1) remain susceptible and increase in age by one year, 2) are infected and move to the corresponding 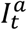 class, or 3) or are immunized by vaccination and move to an immune class (which is not explicitly modeled). Thus, 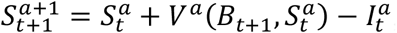, where 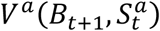 is the impact of vaccination on the susceptible population and is assumed known. For a = 1, this reflects the number of non-immunized births as a function of MCV1 coverage and the efficacy: new susceptibles are new births times (1 – MCV1 coverage x efficacy), where efficacy is assumed to be 84 or 93% for countries that administer the first dose at 9 or 12 months respectively (6,20). For a = 2, this reflects the impact of MCV2. In years with SIAs, this reflects the impact of the SIA on the target age classes. Below we describe how these coverages are combined to reflect alternative assumptions about the correlation among vaccine sources.

The model presented in Simons et al (6) was not age-specific in the first stage. The total number of cases was first estimated using a Kalman filter and then the estimate of total cases was disaggregated to age-specific cases based on an analysis of the distribution of observed cases by WHO region and MCV1 coverage level. Thus, the age distribution of cases for a country did not change from year to year unless the MCV1 level changed from one coverage stratum to another (coverage strata: 0-59%, 60-84%,85-100%). The changes described above result in an age distribution of infections and the susceptible population that changes year-to-year depending on the attack rate and age-targeted vaccination.

The number of individuals in each age class that are infected, 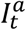, is modeled as binomial draw from the number susceptible with probability *π*_*t*_ that is a logistic function of the proportion susceptible

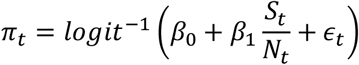

where *N*_*t*_ and *S*_*t*_ are the total and susceptible population at year t, summed over all age cohorts, *β*_0_ and *β*_1_ are unknown parameters to be estimated independently for each country, and *ϵ*_*t*_ is a normal random variate.

The observed number of cases in year t, *C*_*t*_, is assumed to be a binomial random draw from the true number of infections, *I*_*t*_, with probability *p*_*t*_. Following Simons et al 4/11/2022 8:28:00 PM *p*_*t*_, is assumed to be constant in all years with “normal” reporting and assumed to be a higher value, 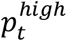, in known years; years with known high reporting rates are specified *a priori* by WHO and are frequently associated with outbreaks or specific activities where surveillance was reinforced. Both *p*_*t*_ and 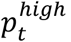 are assumed unknown and are fit independently for each country.

In practice, for computational speed, we approximate the likelihood as a mixture of a normal distribution with mean M and variance V and a Cauchy distribution with mean M and scale V; where M and V are the corresponding mean and variance of the binomial distribution. The mixture is 0.99 weight on the Normal distribution. This approximation is faster computationally because it does not evaluate the large factorials for the equivalent binomial likelihood and the fat tail of the Cauchy prevents complete particle depletion when evaluating parameter combinations with very low likelihood.

### Ensemble of models

In the above model, V_a_(B_t+1_,S_at_), describes the impact of vaccination on the number of susceptible individuals in each age class. Measles vaccination is administered through three mechanisms (routine first dose, routine second dose, and SIAs) that are unlikely to be completely independent (9,10). If the second doses (MCV2 or SIAs) are disproportionately delivered to individuals who have received the first dose (MCV1) and therefore already immune, then the effect of a given coverage level will be reduced relative to the same effect if doses were delivered independently at random (FIGURE 1). Though the second dose coverage is defined as the proportion of first dose recipients who receive the second dose, the explicit correlation of first and second doses is unknown for any country (and likely variable within country), so we represent 4 alternative models (FIGURE 1) that reflect the extreme assumptions that

1. all doses are delivered independently at random: the independent model,
2. MCV2 is disproportionately delivered to recipients of MCV1: the MCV2-MCV1 correlated model,
3. SIA doses are disproportionately delivered to recipients of MCV1: the SIA-MCV1 correlated model,
4. both MCV2 and SIA doses are disproportionately delivered to recipients of MCV1: the all doses correlated model.

**Figure 1.**
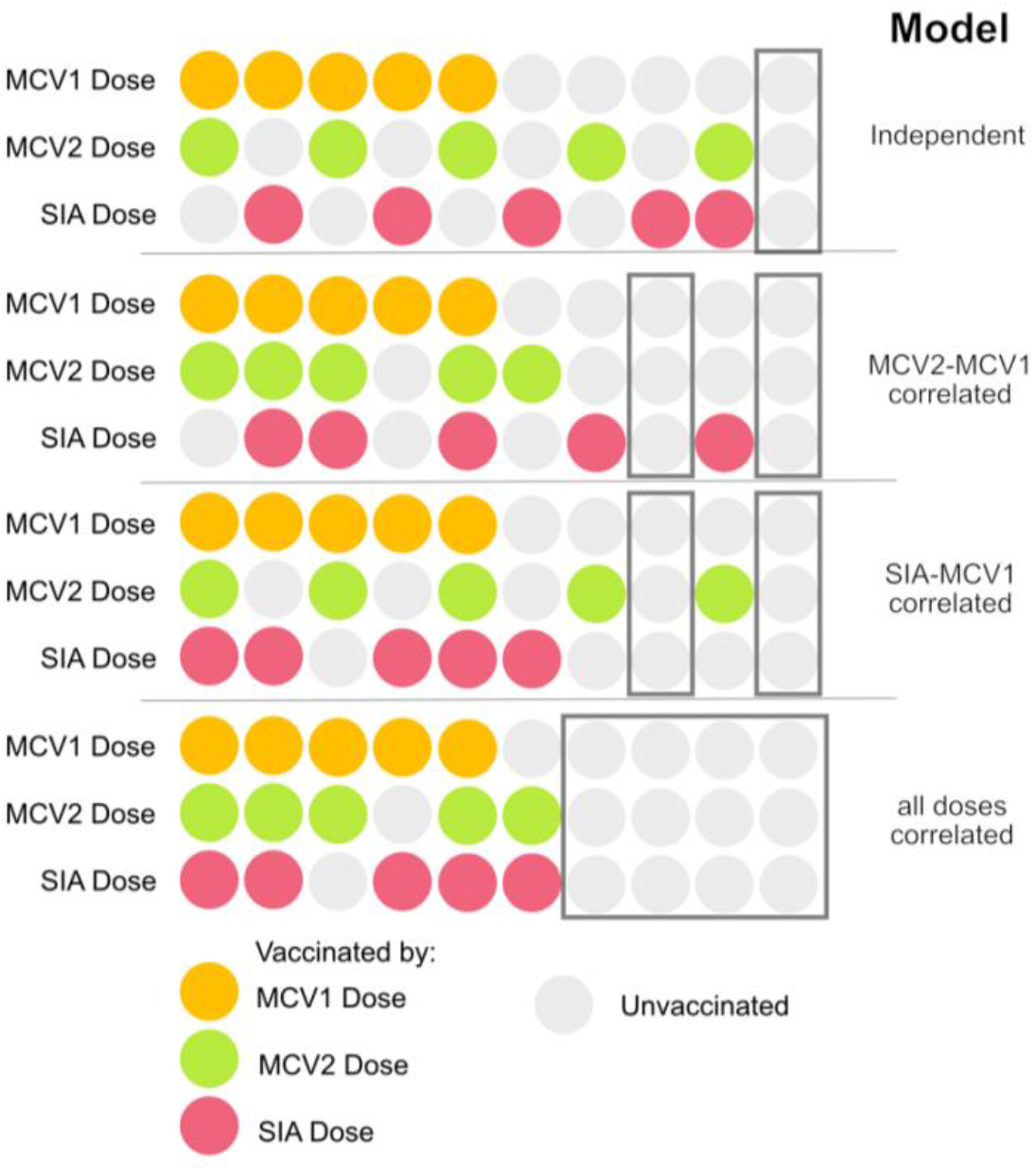
Schematic of 4 alternative models for the interaction of doses of MCV. Here illustrated for a case-study where the coverage for MCV1, MCV2, and SIAs are all 50%. The independent model assumes that all doses of MCV are administered independently. This model is the most optimistic and assumes that 90% of individuals will have receive at least 1 dose. The MCV2-MCV1 correlated and SIA-MCV1 correlated models assume that MCV2 or SIA doses (respectively) are more likely to be received by individuals who have received MCV1. Thus, more doses are expected to be redundant and only 80% of individuals will have received at least 1 dose. The all doses correlated model assumes that MCV2 and SIA doses are more likely to be received by individuals who have received MCV1. This is the most pessimistic scenario and assumes that only 60% of individuals will have received at least one dose.

Specifically, for models 2 and 4 (above) the impact of MCV2 is restricted to only those that received MCV1, P_MCV1_, and thus the effective new proportion of all remaining susceptibles (those that either didn’t receive MCV1 or failed to seroconvert after MCV1) immunized, given MCV2 coverage is P_MCV2_ is

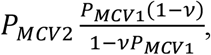

where *υ* is first dose efficacy and the term in the fraction is the proportion of all susceptibles that received the first routine dose but failed to seroconvert. For models 3 and 4 (above) the proportion of all remaining susceptibles that are immunized by SIAs, 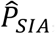, depends whether the SIA coverage, P_SIA_, is greater than the MCV1 coverage, P_MCV1_, as follows:

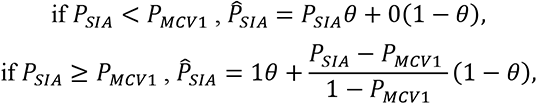

where *θ* = *P*_*MCV*1_(1 − ν)/(1 − ν*P*_*MCV*1_), defined above as the proportion of susceptibles that received MCV1 that failed to seroconvert. Thus, in the first term, the SIA does not reach those who failed to receive MCV1 and in the second term the SIA doses are allocated first to those who received MCV1 and remaining doses are allocated to those who did not.

Depending on the timing of campaigns and the predominant seasonality of measles, the impact of SIAs on case numbers may be observed in the year of the campaign or in the year following the campaign. In practice, this should be considered independently for each SIA; however, the dates of SIAs are not consistently recorded at this level of resolution. Here we fit two versions of each model: one in which the SIA reduces susceptibles in the year in which it was conducted and one in which the SIA reduces susceptibles in the year following the campaign.

### Model fitting and state estimation

For each of the eight candidate models (4 assumptions about dose correlation and 2 assumptions about the timing of SIA impact, above) we evaluate the likelihood using a two-stage grid search of candidate parameter combinations. As described in (19) we search an initial grid of 2000 candidate values for *β*_0_, *β*_1_, *p*_*t*_ and 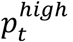. We then conduct a second grid search on a reduced range of the parameter space with another 2000 parameter combinations. This reduced range corresponds to the parameter space that contains the highest 5% of likelihood values in the initial search. At each parameter combination we estimate the likelihood using a particle filter with 1000 particles (19). We take, as our estimate of the likelihood, the highest value of the likelihood from this grid, and the corresponding parameters as the maximum likelihood estimates. For each country, we select the best model as that model with the highest likelihood at its maximum likelihood parameter estimates. In the supplement we illustrate the performance of this likelihood-based model selection using a simulation experiment. Briefly, for 5 country examples, we simulate observed case dynamics under each of several models and then fit all candidate models to these simulated time series. We then quantify the percentage of times for which the generating model is selected as the first or second best-fit.

To estimate the number of unobserved cases, we resample 100 parameter combinations from the grid of possible combinations, with replacement, with probability proportional to the likelihood of each parameter combination. We then evaluate the particle filter at these 100 parameter combinations (many of which may be identical) and combine the resulting distribution of particles. We take the mean and quantiles of this distribution as estimates of the mean and confidence interval for the true number of infections in each age cohort and year.

### Global sums

Following Simons et al, for 93 countries that have good surveillance quality we did not apply the above state-space model. Instead, we estimated the mean number of cases assuming 20% reporting, with lower and upper bounds assuming 40% and 5% reporting respectively (21).

For the remaining countries, we apply the state-space model described above to estimate the number of cases in each one-year age cohort in each year. Though the model is fit to observations from 1980-2019, we only present global sums from 2000-2019 as some countries did not consistently report measles cases prior. We then estimate the corresponding number of measles deaths by multiplying the estimated number of infections by the corresponding country-specific case fatality probability as reported in Wolfson et al (22). As in Simons et al (6) we assumed that the case fatality ratio (CFR) for children above 5 years of age was half that reported in Wolfson et al (22) and 0 for individuals greater than 15 years.

For global totals of measles infections and deaths, we present 2 totals. First, we present an optimistic total, where all countries fitted with model that assumes independent doses; this assumes that all second opportunities are delivered independently and at random with respect the receipt of the first routine dose and is the assumption made in the Simons et al (6) model that has been used for the annual updates (1). Second, we present a total where the estimate for each country comes from the model, selected from the 8 candidates, with the highest likelihood. In the supplement, we present an estimate of total burden where the estimate for each country is the weighted average over all 8 models in the ensemble.

We compare the estimated burden of measles infection and mortality from the state-space model described here to the estimates from the Simons et al 2012 model as reported in (1).

## Results

### Model selection for each country

The best fit model for most countries (78/100) was either the SIA-MCV1 or all doses correlated model; suggesting that the effectiveness of either SIA doses or both SIA and MCV2 doses is lower than the nominal coverage level (FIGURE 2). For 65 countries a single model accounted for >90% of the likelihood weight in the ensemble; for 30 countries, >90% of the likelihood weight was on two models; and only 5 countries required 3 models to account for 90% of likelihood weight (FIGURE 2). For 61 the impact of SIAs was realized as reductions in cases in the year after the SIA rather than in the year of the SIA. On simulated time series, the generating model was correctly selected as the best-fit, or among the top two best-fit model in 80% and 93% of simulations respectively.

**Figure 2.**
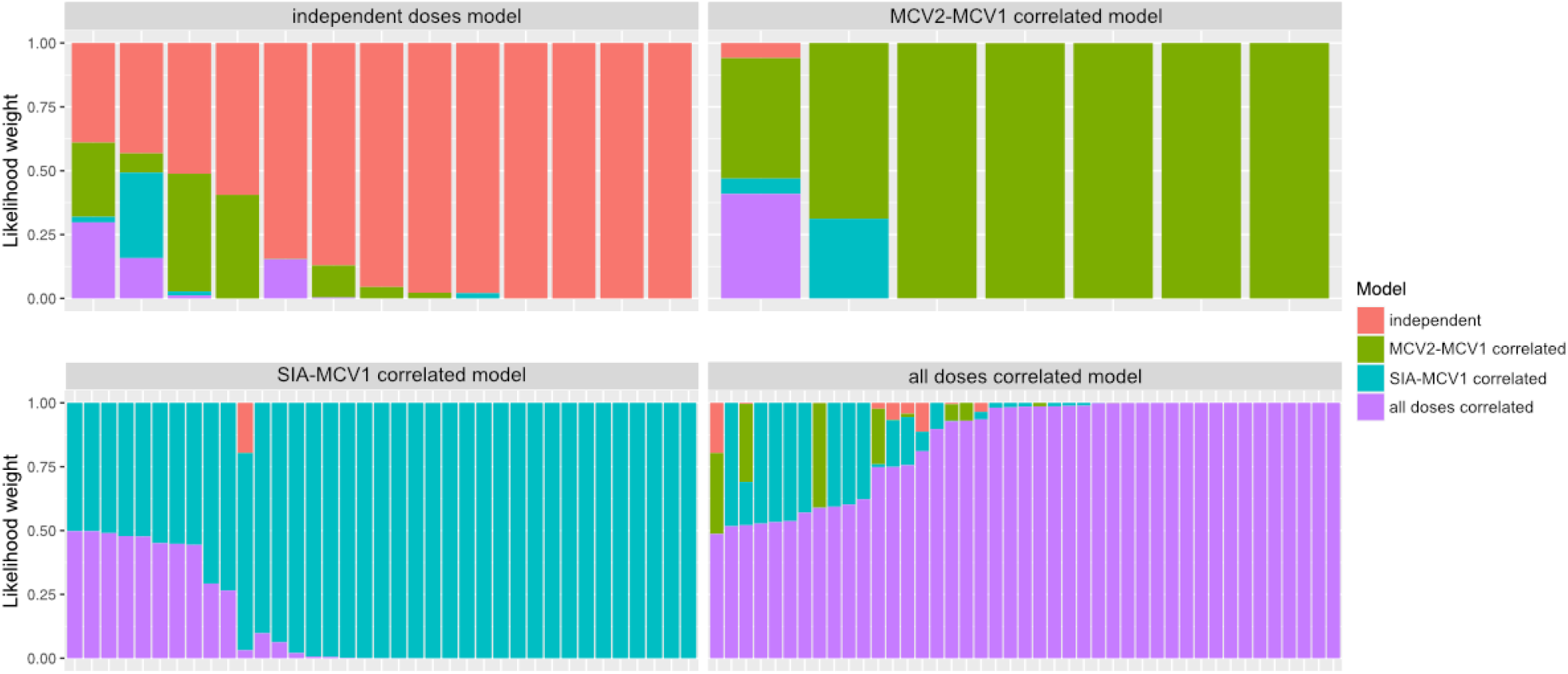
Likelihood weights for each of 4 dose interaction models for each country. Clockwise from top left panels contain countries for which the highest weighted model was the optimistic, MCV2 reduced, SIA reduced, and MCV2 and SIA reduced model respectively.

### Global Burden of Measles

The mean estimate of the total burden of measles infection using the independent doses model is similar to that from the Simons model (6), which makes the same assumptions about the interaction among vaccine doses. The confidence intervals for the new model estimates are considerably reduced, reflecting the different error structure. Notably, because the new model assumes a binomial error, the width of the confidence interval scales with the mean burden of cases.

As expected, the estimates of total burden of measles infection and deaths is lower for the independent doses model, which assumes independence of second dose opportunities, than when the global sum uses the county-specific best-model (FIGURE 3). The model weighted estimate of burden is very similar to the country-specific best-model estimate; most countries are heavily weighted on a single model (Supplementary Material).

**Figure 3.**
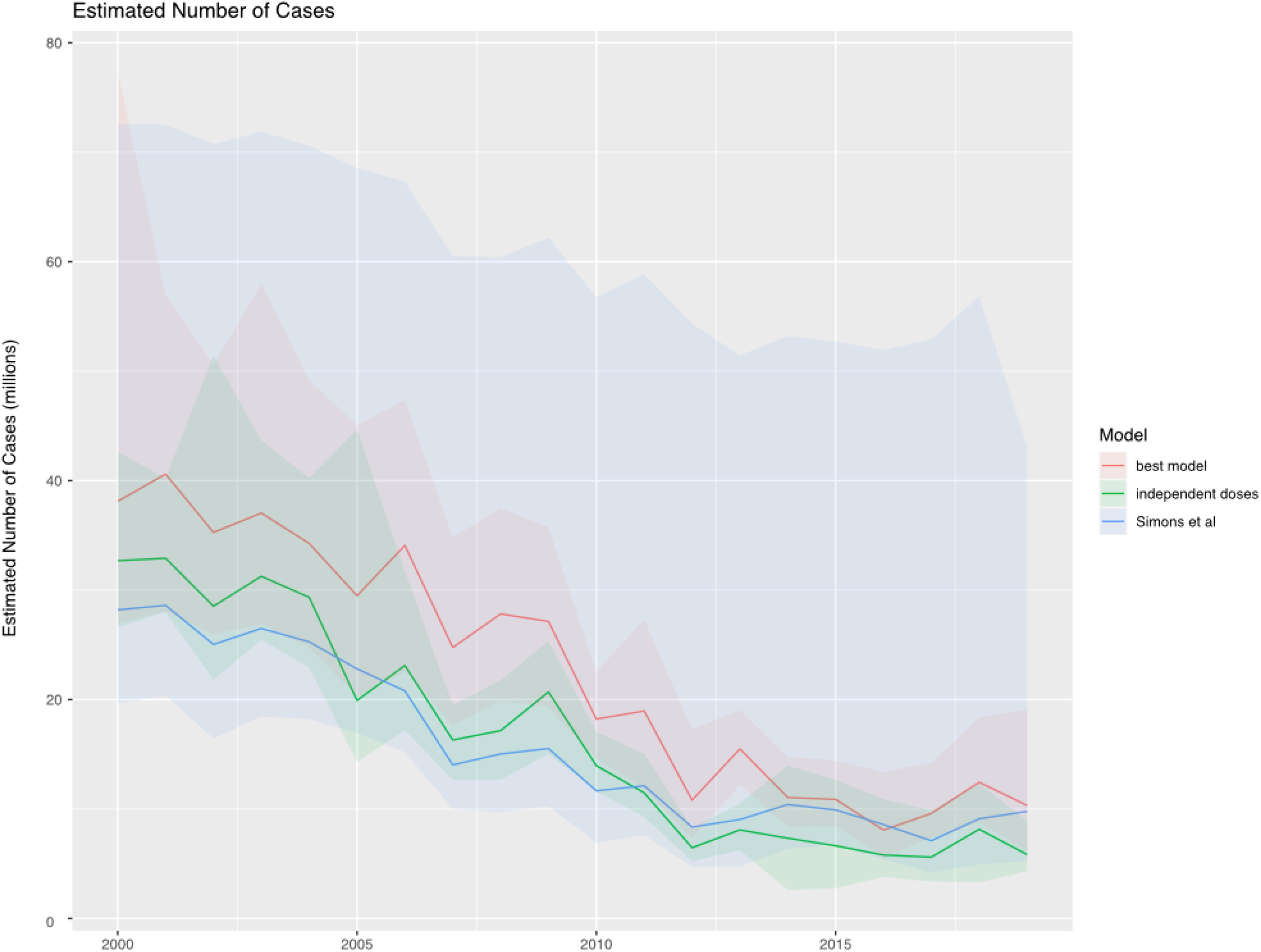
Estimated total burden of measles infection in 193 countries using 3 methods: the country-specific best model (red), the optimistic total assuming independent doses (green), and the Simons et al model (blue). Solid lines indicate the mean estimate and shading indicates the 2.5^th^ and 97.5^th^ percentile.

**Figure 4.**
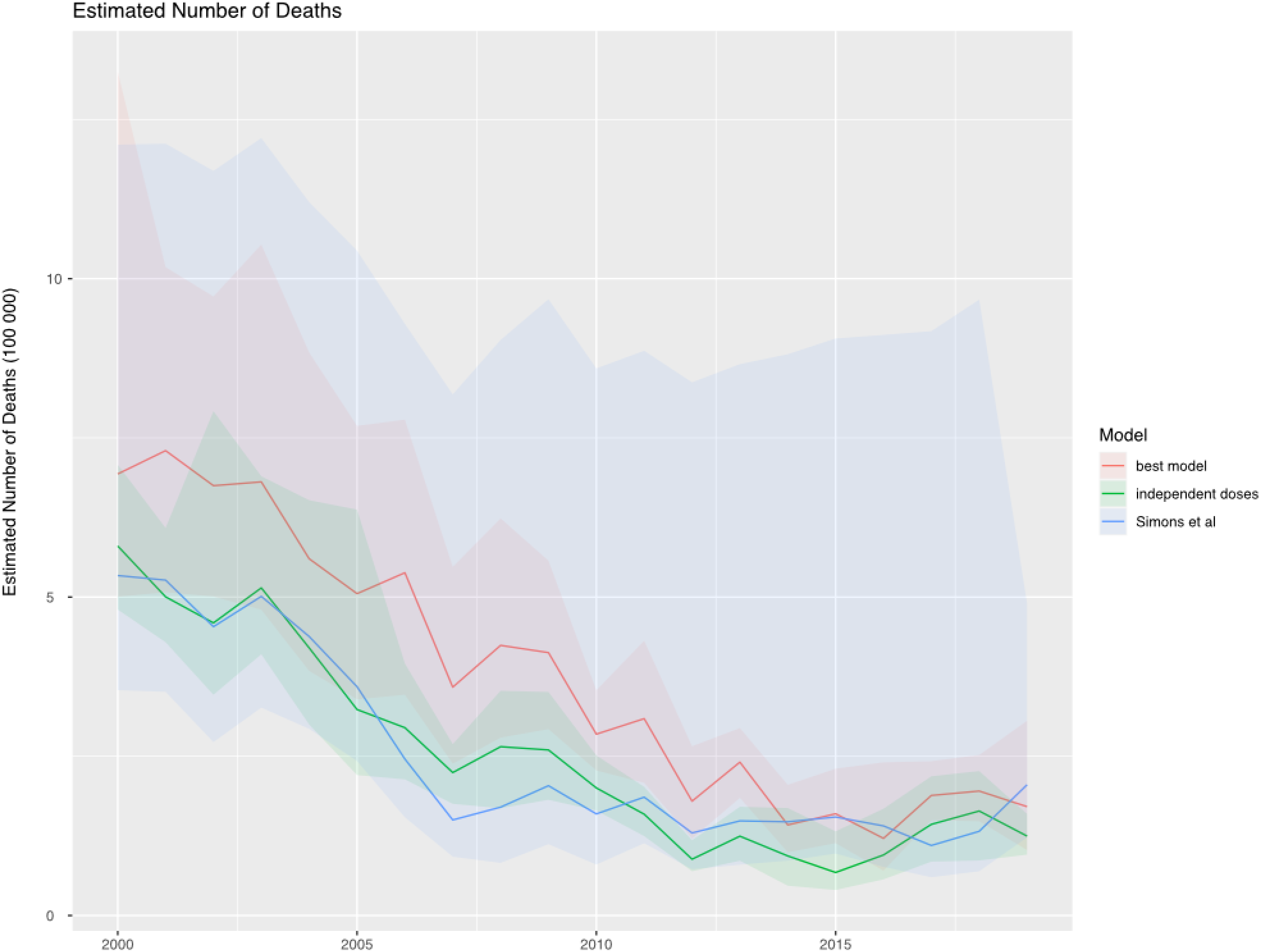
Estimated total burden of measles mortality in 193 countries using 3 methods: the country-specific best model (red), the optimistic total assuming independent doses (green), and the Simons et al model (blue). Solid lines indicate the mean estimate and shading indicates the 2.5^th^ and 97.5^th^ percentile.

Though the country-specific best model estimates are entirely contained within the confidence bounds from the Simons model estimates, the proportional reduction in the mean burden of infection and mortality is greater for the estimates from the new model described here. The country-specific best-model fit estimates higher burden of infection (mean (95% CI): 454 (333-662) million cases between 2000-2019) and mortality (mean (95% CI): 7 (5-11) million deaths between 2000-2019) compared to the Simons model (mean (95% CI): 318 (212-1209) million cases between 2000-2019 and mean (95% CI): 5 (3-19) million deaths). However, the proportional reduction in infections from 2000 to 2019 is greater for the country-specific best-model estimate (73%) compared to the Simons model estimate (65%), because the former estimates higher incidence in 2000 (TABLE 1). Though the impact of SIAs are generally found to perform lower than nominal levels (FIGURE 2), the new age-specific model means that the distribution of cases in each year is more reactive to the impact of age-targeted SIAs. The assumed age distribution in the Simons model changes only in response to routine first-dose MCV coverage. The country-specific best-fit estimates for the new model predict that a smaller fraction of cases are under 5 years of age (e.g. in 2017, 86/100 countries were predicted to have a smaller fraction of cases under 5 years than in the Simons model). Since the CFR is assumed to be higher in children under 5 years (22), this results in an overall decrease in total CFR.

**Table 1.** Total cases of measles and deaths due to measles in 2000 and 2019 estimated by three different methods. Values are the mean estimate summed over 193 countries with 95% confidence intervals in brackets.

| Year | Estimated Cases (millions) |  |  |
| --- | --- | --- | --- |
|  | Best Model | Independent doses | Simons et al |
| 2000 | 38 [27, 72] | 33 [26, 42] | 28 [19, 72] |
| 2019 | 10 [6, 19] | 6 [4, 9] | 10 [5, 43] |

| Year | Estimated Deaths (millions) |  |  |
| --- | --- | --- | --- |
|  | Best Model | Independent doses | Simons et al |
| 2000 | 0.7 [0.5, 1.3] | 0.6 [0.5, 0.7] | 0.5 [0.3, 1.2] |
| 2019 | 0.2 [0.1, 0.3] | 0.1 [0.1, 0.2) | 0.2 [0.1, 0.5) |

Both the country-specific best-model sum and the Simons model estimate an increase in measles infections and deaths from a minimum mean value in 2016 and 2017, respectively. The estimated resurgence of measles infections reflects a 54% (compared to 2018) and 38% (compared to 2019) increase in the country-specific best-model sum and the Simons models, respectively.

## Discussion

Though there have been dramatic reductions in the overall burden of measles disease and mortality over the last 2 decades, the recent resurgence of measles remains a concern. The methods we have described here result in narrower confidence intervals on estimated measles burden, which highlights that the recent increases are unlikely to be the result of random variation in reporting. Estimates derived from the prior method described in Simons et al (6) result in confidence intervals that do not scale with mean incidence and thereby obscure the recent increases in burden.

By fitting an ensemble of models, reflecting alternative assumptions about the correlation between first and second dose opportunities (routine and SIAs) this analysis highlights the dependence of burden estimates on these assumptions. Despite various changes in model structure (see Methods) assuming independence of first and second dose opportunities in the new model results in central estimates of the burden of measles that are consistent with the Simons model estimates. However, we find that the majority of countries were better fit by models that assume that either SIAs or both SIAs and routine second doses are correlated with the first dose.

Though the SIA-MCV1 correlated and all doses correlated models were better supported, we cannot infer directly the reason that the time series for these countries are less well fit by the independent or MCV2-MCV1 correlated models. The formulation of the SIA-MCV1 and all doses correlated models are consistent with a strict interpretation of correlation among first and second dose opportunities; specifically, that second doses are delivered first to those who have received a prior first dose and remaining doses are then randomly distributed to those who have not received a first dose. Phenomenologically, this results in dynamics that are consistent with routine second dose and SIA coverage that is lower than the nominal administrative coverage. This difference could arise because of dependence among doses, which could be due to inequity in access to both routine and SIA services (23,24). Alternatively, this same pattern could arise if administrative SIA coverage routinely overestimates coverage achieved (e.g. because of underestimates of the target population size). We note that the 4 candidate models we explore here are extreme assumptions of either no correlation or strict correlation among doses. The truth is likely to be intermediate in all cases and future work should explore support for alternative models.

Regardless of mechanism, the finding that observed dynamics are frequently consistent with routine second doses and SIAs performing below nominal coverage has important implications for planning of measles vaccination investments. Countries for which second dose opportunities that underperform nominal coverage may be expected to continue to do so in the future in the absence of explicit investments to address equity in coverage. Here we fit an ensemble of models that range from optimistic (independent) to pessimistic (all doses correlated) assumptions about the immunization impact of second dose programs. Future projections of the impact of vaccination investments could use this ensemble of models, with equal weights or weights based on likelihood (25), to reflect uncertainty about the future performance of vaccine investments.

Ideally, the findings from this modeling exercise should be corroborated with studies of the correlation between vaccine opportunities in select countries.

As in the method described in Simons et al (6), this model does not include several important aspects of measles transmission dynamics because of limited data availability over the full time series. Specifically, we ignore the effect of seasonal variation in measles transmission because consistent data are only available at the annual time scale for most countries since 1980. Further, we assume homogenous mixing among all age classes because age-specific case reporting is not consistently available over this time period. Efforts to develop age-specific mixing matrices for all countries (e.g.(26)) only consider recent time and would require additional strong, but untestable, assumptions about the structure of these matrices over time. Thus, the role of age-structure in this model is to permit explicit representation of age-targeted vaccination interventions. The availability of weekly or monthly and age-specific case reporting (e.g. (2)) could permit the incorporation of these dynamics in the future but would necessarily come at the cost of shorter (though more highly resolved) time series. Finally, we assume that vaccination coverage, due to all sources, is homogeneous at the sub-national level. Various analyses have shown that subnational variation in vaccination coverage can be significant (24,27–29) and meaningfully affect transmission dynamics (30). Methods to reconcile fine-scale assessment of vaccination coverage with aggregate national or sub-national case reporting remains an important area of research.

This work highlights the importance of continuing to update models for burden estimation to account for technical and data quality advances. Here we show that the existing methods, based on an optimistic assumption of the impact of second dose opportunities lead to lower estimates of the burden of measles disease and mortality and are less well supported when compared to models that assume that receipt of the second dose is correlated with the receipt of the first dose. The performance of measles control and elimination would benefit from both improved coverage and efforts to extend vaccination to populations consistently missed by current programs (24,31). Further, investment in strengthening national surveillance programs would permit continued development of mechanistic burden estimation models that can account for dynamics (age-structured mixing, seasonality) that must currently be excluded due to limitations of data quality.

## Data Availability

The data referred to this manuscript are available at the websites referenced in the text.

https://apps.who.int/immunization_monitoring/globalsummary/timeseries/tsincidencemeasles.html

https://population.un.org/wpp/

https://cdn.who.int/media/docs/default-source/immunization/data_statistics/summary_measles_sias.xls?sfvrsn=77c794e8_6

