## Supplemental Material for "Ensemble model estimates of the global burden of measles morbidity and mortality from 2000 to 2019: a modeling study"

*S1. Model Weighted Estimates*

To generate model-weighted estimates of the number of true infections we take a model weighted average of the mean, $2.5^{th}$ , and 97.5th quantiles of the estimated cases in age group a at time t:

$$\hat{I}_t^a = \sum_{m=1}^{8} p_m I_{t,m}^a$$

where $p_m = \frac{l_m}{\sum_x l_x}$ is the normalized likelihood weight of the maximum likelihood, $l_m$, for each model in the ensemble. Here $I_{t,m}^a$ is the corresponding summary statistic (mean or quartile) of the distribution of particles in age group a at time t. We generated model-weighted summaries for each of the 100 countries fit with the particle filter. We combined these with the remaining countries to generate a global estimate (Figure S1) as described in the main text.

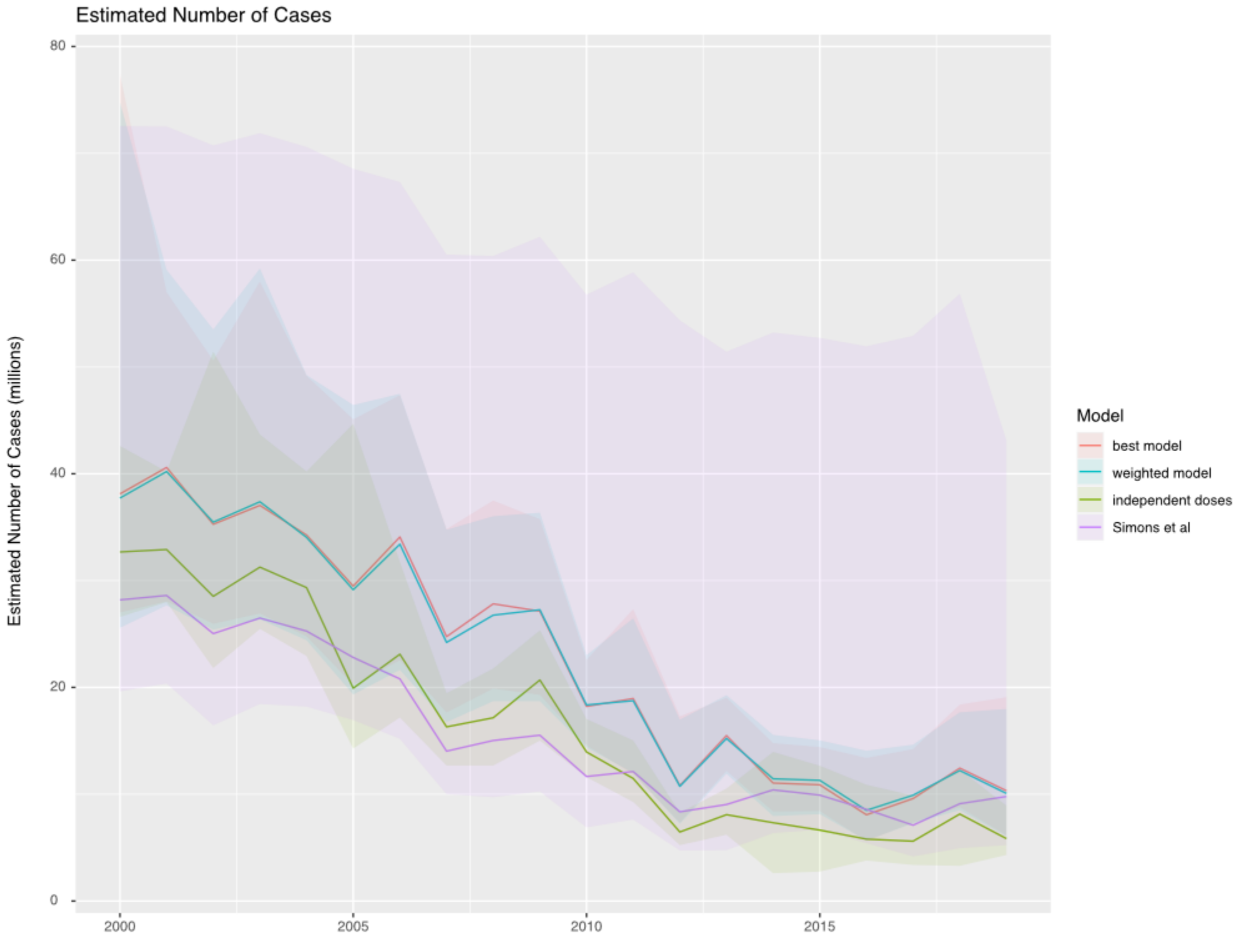


*Figure S1. Estimated total burden of measles infection in 193 countries using 4 methods: the country-specific best model (red), the country-specific model-weighted average (blue), the optimistic total assuming independent doses (green), and the Simons et al model (purple). Solid lines indicate the mean estimate and shading indicates the 2.5$^{th}$ and 97.5$^{th}$ percentile.*

### *S1. Model Selection Simulation Study*

To verify the ability of our likelihood-based model selection to identify the correct model, we conducted the following simulation study. First, we selected 5 countries with high measles incidence, fit all candidate models, and obtained estimates of $\beta_0$ and $\beta_1$. These countries (which will not be identified) were chosen because they reflect a large fraction of global measles burden and they reflect a variety of historical vaccination patterns; all had at least one SIA, though some had many, and some have not yet introduced a routine second vaccine dose (MCV2; Table S1). Further these countries reflect a range of parameter estimates for $\beta_0$ and $\beta_1$. We then used the fitted parameter values to generate multiple simulated time series of infected individuals from 1980-2020 under each of several of the candidate models (Table S1); for each country we included one simulation for which the impact of SIAs is realized in the year following the SIA ("SIAs shifted" in Table S1).  Note that for countries that had not introduced MCV2 we did not generate forward simulations under the MCV2 model (Table S1). For each, we used the corresponding population size, birth rate, and vaccination history for the country. We then simulated the reported cases in each country as a binomial draw from the simulated infected individuals with reporting probability 0.01.

We then fit all 8 of the candidate models from the main text to the simulated time series for each country and model combination. The results of model selection for each country are illustrated in Tables S2-6. For 4 of the 5 country simulations, the generating model was selected was the best fit (highest estimated log likelihood) in >50% of simulated time series. For 11 out of 19 country-model combinations, the generating model was in the top 2 ranked models in 100% of simulated time series, and for 14 out of 19 country-model combinations the generating model was in the top 2 ranked models in at least 90% of simulated time series. The cases where the performance of model selection were poor were restricted to a single country setting, suggesting that there may be demographic or vaccine history settings where model selection performs less well.

Table S1. The parameter values and generating model combinations for the 5 country scenarios simulated for the model selection study. Dots indicate county-generating models that were simulated.

| | | Country | | | | |
|---|---|---|---|---|---|---|
| | Parameter | A | B | C | D | E |
| | $\beta_0$ | -4.4 | -6.0 | -4.3 | -4.4 | -4.3 |
| | $\beta_1$ | 21 | 41 | 23.1 | 119 | 105 |
| Generating Model | Independent | ● | ● | ● | ● | ● |
| | MCV2-MCV1 Correlated | | | | ● | ● |
| | SIA-MCV1 Correlated | ● | ● | ● | ● | ● |
| | All Doses Correlated | | | | ● | ● |
| | Independent, SIAs shifted | ● | ● | ● | ● | ● |

Table S2: Model fitting results for simulated time series using Country A settings.

| Generating Model | # simulations | % simulations where generating model was ranked first | % simulations where generating model was ranked first or second |
|---|---|---|---|
| Independent | 33 | 91% (30/33) | 100% (33/33) |
| SIA-MCV1 Correlated | 33 | 73% (24/33) | 91% (30/33) |
| Independent, SIAs shifted | 33 | 100% (33/33) | 100% (33/33) |
| All models | 99 | 88% (87/99) | 97% (97/99) |

Table S3: Model fitting results for simulated time series using Country B settings.

| Generating Model | # simulations | % simulations where generating model was ranked first | % simulations where generating model was ranked first or second |
|---|---|---|---|
| Independent | 33 | 100% (33/33) | 100% (33/33) |
| SIA-MCV1 Correlated | 33 | 88% (29/33) | 100% (33/33) |
| Independent, SIAs shifted | 33 | 100% (33/33) | 100% (33/33) |
| All models | 99 | 96% (95/99) | 100% (99/99) |

Table S4: Model fitting results for simulated time series using Country C settings.

| Generating Model | # simulations | % simulations where generating model was ranked first | % simulations where generating model was ranked first or second |
|---|---|---|---|
| Independent | 33 | 100% (30/33) | 100% (33/33) |
| SIA-MCV1 Correlated | 33 | 85% (28/33) | 100% (33/33) |
| Independent, SIAs shifted | 33 | 97% (32/33) | 100% (33/33) |
| All models | 99 | 91% (90/99) | 100% (99/99) |

Table S5: Model fitting results for simulated time series using Country D settings.

| Generating Model | # simulations | % simulations where generating model was ranked first | % simulations where generating model was ranked first or second |
|---|---|---|---|
| Independent | 20 | 55% (11/20) | 70% (14/20) |
| MCV-MCV1 Correlated | 20 | 60% (12/20) | 80% (16/20) |
| SIA-MCV1 Correlated | 20 | 45% (9/20) | 60% (12/20) |
| All Doses Correlated | 20 | 40% (8/20) | 70% (14/20) |
| Independent, SIAs shifted | 20 | 50% (10/20) | 80% (16/20) |
| All models | 100 | 50% (50/100) | 72% (72/100) |

Table S6: Model fitting results for simulated time series using Country E settings.

| Generating Model | # simulations | % simulations where generating model was ranked first | % simulations where generating model was ranked first or second |
|---|---|---|---|
| Independent | 20 | 80% (16/20) | 100% (20/20) |
| MCV-MCV1 Correlated | 20 | 75% (15/20) | 100% (20/20) |
| SIA-MCV1 Correlated | 20 | 70% (14/20) | 95% (19/20) |
| All Doses Correlated | 20 | 80% (16/20) | 100% (20/20) |
| Independent, SIAs shifted | 20 | 85% (17/20) | 90% (18/20) |
| All models | 100 | 78% (78100) | 97% (97/100) |
